# Is *SORL1* a common genetic target across neurodegenerative diseases?: A multi-ancestry biobank scale assessment

**DOI:** 10.64898/2026.01.24.26344530

**Authors:** Marzieh Khani, Sheila N. Yeboah, Catalina Cerquera-Cleves, Alexandra Kedmi, Bernabe I. Bustos, Spencer M. Grant, Suleyman Can Akerman, Fulya Akçimen, Paul Suhwan Lee, Paula Reyes-Pérez, Lara M. Lange, Hampton Leonard, Mathew J. Koretsky, Mary B. Makarious, Zachary Schneider, Caroline Jonson, Pin-Shiuan Chen, Yi Wen Tay, Jeffrey D Rothstein, Chin-Hsien Lin, Shen-Yang Lim, Christine Klein, Kalpana Merchant, Niccolò E Mencacci, Dimitri Krainc, Mark R. Cookson, Global Parkinson’s Genetics Program (GP2), Andrew Singleton, Sara Bandres-Ciga

## Abstract

*SORL1,* the gene encoding the SORLA protein, has arisen as a potential therapeutic target for Alzheimer’s disease (AD). Studies suggest that restoring SORLA function or its trafficking pathways, particularly the SORLA–retromer recycling system, may offer a promising strategy to slow or halt AD progression. While both rare and common *SORL1* variants have been associated with increased AD risk, recent evidence suggests a potential involvement of *SORL1* in other neurodegenerative conditions. This study assessed the contribution of *SORL1* genetic variation to the risk of AD, related dementias (RD), and Parkinson’s disease (PD) using data from six large-scale biobanks, comprising 15,043 AD, 9,943 RD, and 42,763 PD cases, along with 111,969 controls across 11 ancestries. We identified 53 potentially disease-related *SORL1* variants (CADD score > 20, MAC ≥ 2, annotated as protein-altering or splicing, and with the mutated allele present only in cases), including 41 novel and 12 previously reported variants. Three were found across multiple ancestries. Overall, 13 variants were found in AD-related cohorts, 5 in RD cohorts, and 35 in PD cohorts. Association analysis identified 10 nominally significant variants associated with AD and 5 with PD. The replication of multiple *SORL1* variants across neurodegenerative diseases and ancestrally diverse populations underscores its potential broad genetic contribution to neurodegeneration and reinforces its relevance across distinct clinical phenotypes. Gene-based burden analysis did not reveal any significant cumulative effect of *SORL1* variants in the populations tested. A family-based analysis identified a rare predicted-damaging variant in two East Asian families (11:121478242:G:A, p.R176Q) and two variants in two families of European ancestry (11:121514222:A:C, p.N371T; 11:121545392:G:A, p.V672M) that show some evidence of segregation in PD families. Although these variants were slightly more frequent in unrelated PD cases vs. controls, none of them showed statistically significant enrichment in PD, likely due to their very low frequency. Overall, our results extend the understanding of *SORL1* beyond AD, suggesting a broader role in neurodegeneration and emphasizing the need for diverse population studies when evaluating genetic risk.

## Introduction

The sortilin-related receptor gene (*SORL1*) encodes the SORLA protein, which is responsible for directing the organization and placement of proteins within cellular compartments. Specifically, SORLA helps regulate the exocytosis of the amyloid precursor protein (APP), thus preventing cleavages in APP that may be potentially pathogenic, and is known to be involved in Alzheimer’s disease (AD) pathology [1,2]. More than 500 *SORL1* protein-altering variants associated with AD have been identified across various ancestries [3–6]. These variants span the etiological risk spectrum, ranging from rare, highly deleterious pathogenic variants to common variants conferring low to moderate risk. About 2.75% of patients with early-onset AD (EOAD) and 1.5% of late-onset AD (LOAD) carry potentially pathogenic *SORL1* variants segregating with an autosomal dominant pattern of inheritance [7]. Deficits in the SORLA-retromer complex are strongly linked to AD pathogenesis. SORLA, an endosomal receptor, partners with the retromer trimer core complex (VPS26-VPS35-VPS29) to regulate the recycling of AD-related cargos like APP. Disruption of SORLA or retromer in models mimics key AD biomarkers and pathology [8]. Restoring this recycling pathway is a promising therapeutic strategy, and ongoing drug development efforts, including small molecules, gene therapy, and integrated biomarker approaches, aim to advance treatments for AD based on this mechanism [9].

In AD patients, most reported *SORL1* variants are rare and missense [10]. A study conducted on a cohort of 1,255 EOAD patients and 1,938 controls from Western Europe identified 44 rare *SORL1* missense variants whose mutated allele was present only in patients, with 80% predicted to be likely pathogenic using in silico tools [11]. Recent research on individuals of Italian ancestry identified three rare missense variants, one of which was declared to be likely pathogenic for EOAD with an autosomal dominant inheritance pattern [12].

The *SORL1* locus has been linked to sporadic LOAD through previous genome-wide association studies (GWAS) [10,12,13]. Recent multi-ancestry GWAS have highlighted the complexity of this locus, suggesting highly heterogeneous effects across populations [14–16]. Common variability at this locus is linked to disease risk in both East Asian and European populations, but despite overlapping signals the underlying genetic architecture appears to be distinct between these groups, or at least not fully resolved [14,15,17].

Of note, *SORL1* variants have also been reported to be putatively linked to Parkinson’s disease (PD), including Parkinson’s disease dementia (PDD). Recently, we reported a family of Greek ancestry identified a heterozygous variant in the *SORL1* gene (p.G379W) linked to autosomal dominant PD and dementia with incomplete penetrance. These findings suggest that *SORL1* variants are also involved in a broader class of neurodegenerative diseases in addition to AD [18]. Research on the role of *SORL1* in PD risk has very recently begun to expand across non-European ancestries. The first study of *SORL1* and its link with PD in Chinese populations found two common variants (rs1010159 and rs2298813) that demonstrated an association with a higher predisposition to PD as compared to controls [19].

This study comprehensively assesses the role of *SORL1* genetic variation in AD, related dementias (RD), and PD, using the largest multi-ancestry genetic datasets available to date, including All of Us (AoU), the Alzheimer’s Disease Sequencing Project (ADSP), Accelerating Medicines Partnership in Parkinson’s Disease (AMP PD), the 100,000 Genomes Project (100KGP), the United Kingdom Biobank (UKB), and the Global Parkinson’s Genetics Program (GP2). Our analysis includes 15,043 AD, 9,943 RD, and 42,763 PD cases, along with 111,969 controls, from 11 genetically determined ancestries. This research investigates a promising therapeutic target across the neurodegenerative disease spectrum and within a global context, representing an essential step towards the new era of precision medicine.

## Methods

### Cohorts under study

AD and RD (defined as anyone with dementia who does not have AD or PD) cohorts included whole-genome sequencing (WGS) data from five data resources: AoU (v7), ADSP (v4), AMP PD (v3), 100KGP (v18)[20], and UKB (v18.1) **(Figure 1)**. PD cohorts included WGS data from AMP PD (v3), AoU (v7), UKB (v18.1), and 100KGP (v18), as well as imputed genotyping data from GP2 (release 9) **(Figure 1)**. Family-based analysis was performed on WGS data in GP2 (release 10) **(Figure 1)**. All samples in AoU, UKB, ADSP, AMP PD, and GP2 underwent quality control and a custom ancestry prediction pipeline included in the GenoTools package (https://github.com/dvitale199/GenoTools). Related individuals, up to second degree relatives, were removed (KINSHIP > 0.0884). Ancestries represented include European (EUR), African (AFR), American Admixed (AMR), African Admixed (AAC), Ashkenazi Jewish (AJ), Central Asian (CAS), Eastern Asian (EAS), South Asian (SAS), Middle Eastern (MDE), Finnish (FIN), and Complex Admixture History (CAH). Additional details on each database related to ADRDs, as well as a more thorough description of the cohort creation, quality control, variant filtering, and analysis processes, are described elsewhere [21].

**Figure 1.**
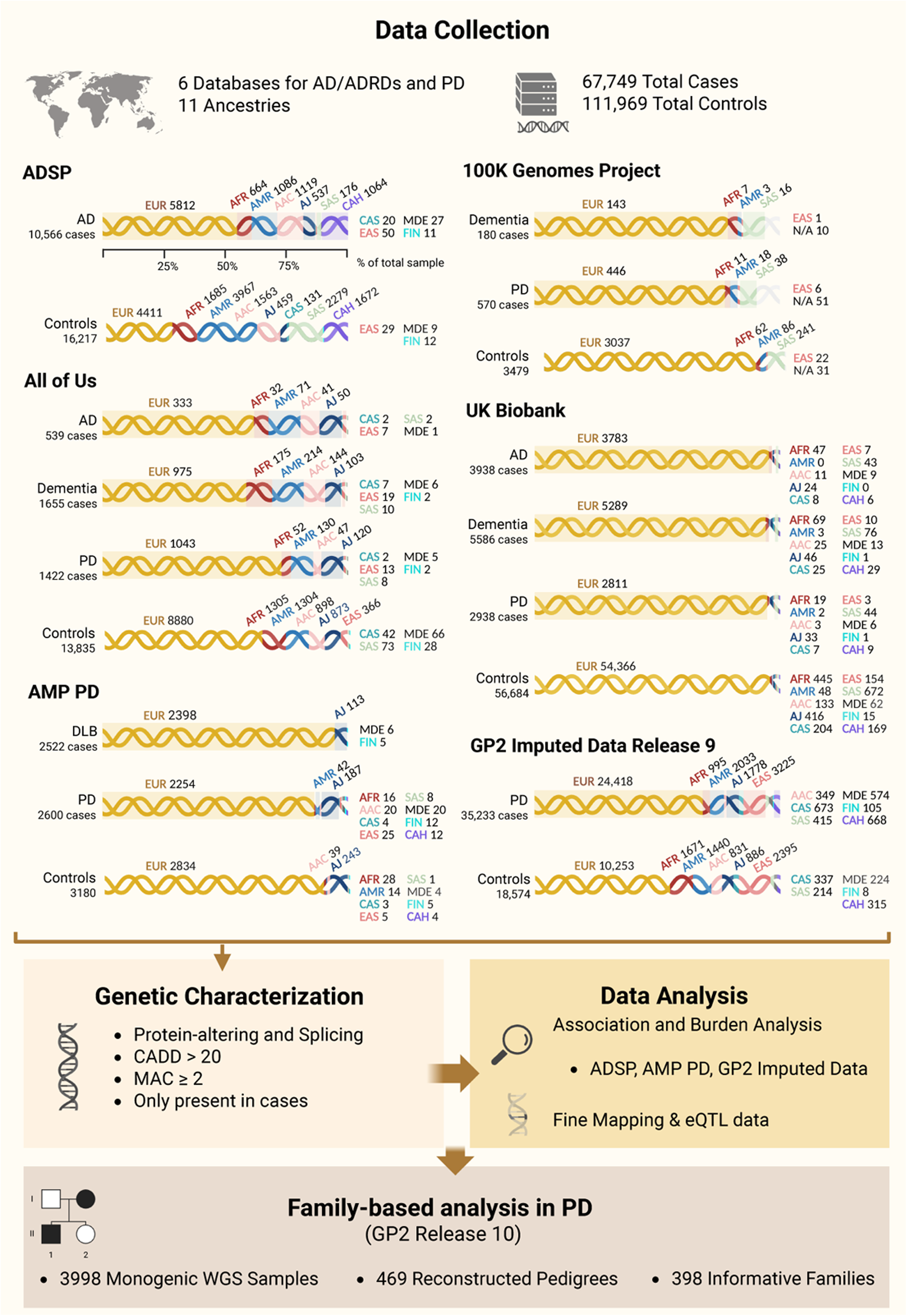
Workflow and demographic characteristics of biobank-scale cohorts under study. The figure illustrates the workflow and the number of cases and controls per ancestry across six datasets in this study: Alzheimer’s Disease Sequencing Project (ADSP), All of Us (AoU), Accelerating Medicines Partnership in Parkinson’s Disease (AMP PD), 100,000 Genomes Project (100KGP), UK Biobank (UKB), and Global Parkinson’s Genetics Program (GP2) Imputed Data. Ancestries represented include European (EUR), African (AFR), American Admixed (AMR), African Admixed (AAC), Ashkenazi Jewish (AJ), Central Asian (CAS), Eastern Asian (EAS), South Asian (SAS), Middle Eastern (MDE), Finnish (FIN), and Complex Admixture History (CAH). N/A, Not Available; AD, Alzheimer’s disease; DLB, Dementia with Lewy Bodies; PD, Parkinson’s disease.

### Genetic characterization

We used PLINK 1.9 [22] and PLINK 2 [23] to extract allele frequencies and explore zygosity in both cases and controls. Functional annotation of *SORL1* variants was done using ANNOVAR [24].

Potential disease-causing variants were filtered to include only those present in cases, with a combined annotation-dependent depletion (CADD) score > 20 (predicted to be among the top 1% most pathogenic in the genome)[25], a minor allele count (MAC) ≥ 2 (to be conservative on possible artifacts), and annotated as protein-altering or splicing. Logistic regression and burden analyses, adjusted for sex, age, and principal components (PCs, 1-10) to account for population stratification, were performed in the most well-powered database for each disease (ADSP for AD, and AMP PD and GP2 imputed data for PD). Burden analyses, adjusted for sex, age, and PCs (1–10), were conducted using RVTESTS v2.1.0 [26] following the optimized nonburden Sequence Kernel Association Test (SKAT-O) method. Association and burden analyses were conducted by functional category, including only exonic and splicing variants with MAC ≥ 2. Synonymous variants were retained for both AD and PD group analyses, as they have previously been associated with AD. In ADSP, age at onset for cases and age at recruitment for controls were used as covariates. In AMP PD and GP2 genotyping imputed data, baseline age, and age at sample collection respectively, were used as covariates for both cases and controls due to extensive missing data on age at onset for cases. In AMP PD, the analyses were conducted in EUR and AJ cohorts only due to sample size constraints for other ancestries **(Figure 1)**.

### Family-based analysis

We performed a family-based analysis using the Monogenic cohort (study_type = “Monogenic”), yielding 3,998 WGS-genotyped individuals. Core pedigrees from the GP2 Monogenic Working Group were combined with kinship-inferred families. Briefly, kinship-inferred families were constructed from the ancestry-specific WGS “.related” files by restricting to Monogenic WGS individuals not already present in the core pedigrees. Within each ancestry, we built a graph of related pairs and defined each connected component (≥2 individuals) as a family. Because relationship direction is not inferred from kinship values alone, members were represented as founders in the resulting PED file. The core and kinship-inferred pedigrees were then merged into a unified family set, and “informative” families were defined as having at least one PD case, and a minimum of two individuals. We used PLINK (version 2.0) [23] to extract informative family members and the *SORL1* locus (GRCh38 chr11:121.45–121.63 Mb) from the joint-called WGS data. Variants in *SORL1* were annotated with ANNOVAR [24] to identify rare, predicted-damaging alleles (loss-of-function (LOF) and missense variants with CADD > 20) [25] with ancestry-specific allele frequency < 1% in gnomAD v4.1[27]. We also used ClinVar within ANNOVAR to identify samples carrying pathogenic/likely pathogenic variants in both causal and high-risk PD genes such as *GBA1*, *LRRK2*, *SNCA*, *VPS35*, *PRKN*, *PINK1*, *PARK7*, *RAB39B* and *RAB32*. For each family per ancestry, we calculated *SORL1* carrier counts, generated variant segregation summaries, and prioritized variants that segregated in one or more families and were present in all genotyped PD members and absent from controls, if controls were present. Finally, we evaluated the prioritized *SORL1* variants in an unrelated GP2 WGS case–control cohort of 10,261 individuals, estimating carrier frequencies in ancestry-specific PD cases and healthy controls, and testing for enrichment in PD within each ancestry using one-sided Fisher’s exact tests in R.

### Fine-Mapping analyses

To perform fine-mapping, we aggregated multiple publicly available GWAS datasets across diverse populations. This included the most recent ADRD GWAS comprising 39,106 clinically diagnosed AD cases, 46,828 proxy-RD cases (defined as unaffected individuals with a parent diagnosed with AD or dementia), and 401,577 controls of European ancestry [3]. We also included FinnGen Release 12, which features 10,821 AD cases and 485,932 neurologically healthy controls for further validation [28].

Ancestry-specific AD datasets were incorporated, including the largest available GWAS of African Americans (2,748 cases and 5,222 controls) [29], East Asians (3,962 cases and 4,074 controls) [30], and Caribbean Hispanics (1,095 cases and 1,179 controls). Additional summary statistics were included from GWAS of AD (744 cases, 19,648 controls), ADRD (4,073 cases, 19,648 controls), and a meta-analysis including ADRD, paternal, and maternal proxy dementia cases from the Million Veteran Program (MVP), combined with Alzheimer’s Disease Genetics Consortium (ADGC) samples from African American ancestry (13,441 cases, 69,627 controls). Finally, a GWAS of 38,130 paternal and 39,797 maternal proxy dementia individuals of African ancestry were also used [31]. For PD, we relied on the latest and largest European study of PD genetic risk to date, with 63,555 cases, 17,700 proxy cases with a family history of PD, and 1,746,386 controls [32].

Fine-mapping was performed using the coloc package (version 6.0.0) in R (version 4.3.1), and a posterior probability (PP) threshold of > 0.6 was applied.

### Functional annotation of Alzheimer’s and Parkinson’s disease-associated variants using multivariate brain eQTL data

We extracted two previously fine-mapped variants in EUR and EAS populations (rs11218343 and rs117807585) [16], along with all variants identified in the association analysis we performed in this study, from the multi-variate multiple QTL (mmQTL) dataset by Zeng et al [33]. The study developed the mmQTL pipeline and applied it to brain tissue data from PsychENCODE, ROSMAP, and GTEx version 8. The pipeline uses linear mixed models for eQTL detection, identifies conditionally independent eQTLs, and combines results across datasets using a random-effects meta-analysis that accounts for correlations across brain regions.

## 3. Results

### Demographic and clinical characteristics of variant carriers across cohorts under study

We identified 33 AD patients, 11 RD patients, and 96 PD patients carrying potentially disease-causing *SORL1* variants across the studied cohorts. Among carriers with available data, 18.18% of AD patients, 9.09% of RD patients, and 27.08% of PD patients reported a family history of AD or PD. In terms of sex distribution, 60.61% of AD patients were female and 39.39% were male; 45.45% of RD patients were female and 54.55% were male; and 39.58% of PD patients were female and 60.42% were male. The average age was 77.40 ± 4.88 years for AD patients, 76.45 ± 6.46 years for RD patients, and 63.81 ± 11.46 years for PD patients. The average age at onset (AAO) was 69.45 ± 9.71 years for AD patients, 76.18 ± 6.49 years for RD patients, and 58.21 ± 12.19 years for PD patients.

### SORL1 variants in Alzheimer’s disease and related dementias

Our analysis included two approaches. First, we examined rare, highly penetrant pathogenic variants that were only present in affected individuals and had the potential to cause disease. Within the ADSP cohort, a total of 10,566 AD cases and 16,217 controls were analyzed (**Figure 1**). Functional annotations of all identified *SORL1* variants by cohort and ancestry in AD and RD before filtering are reported in **Supplementary Table 1**. After filtering, 10 potential disease-causing variants that passed our criteria (MAC ≥ 2, CADD > 20, protein-altering or splicing, and present only in cases) remained — eight in EUR (11:121514282:A:G[p.Y391C], 11:121570255:G:A[p.D1108N], 11:121545246:T:C[p.V623A], 11:121559561:C:T[p.R985X], 11:121589355:A:G[p.D1348G], 11:121618787:T:A[p.F1873Y], 11:121625182:G:T[p.G2090V], 11:121627701:G:A[p.A2171T]) and two in AAC (11:121566979:G:-[p.C1030fs], 11:121570183:C:T[p.R1084C]) ancestries. Of these, three (11:121514282:A:G[p.Y391C], 11:121625182:G:T[p.G2090V], 11:121570183:C:T[p.R1084C]) were previously reported in AD [11,34,35], while seven were novel. Identified variants, zygosity, and allele frequencies for each ancestry are reported in **Table 1**. The 11:121514282:A:G[p.Y391C] variant was also observed in one AD patient of AAC ancestry. Family history was not reported for any of these individuals. Among the 23 carriers, 14 were classified as LOAD (60.9%, AAO ≥65 years) and 9 as EOAD (39.1%). Clinical and neuropathological characteristics for all carriers are reported in **Supplementary Table 2**.

**Table 1.**
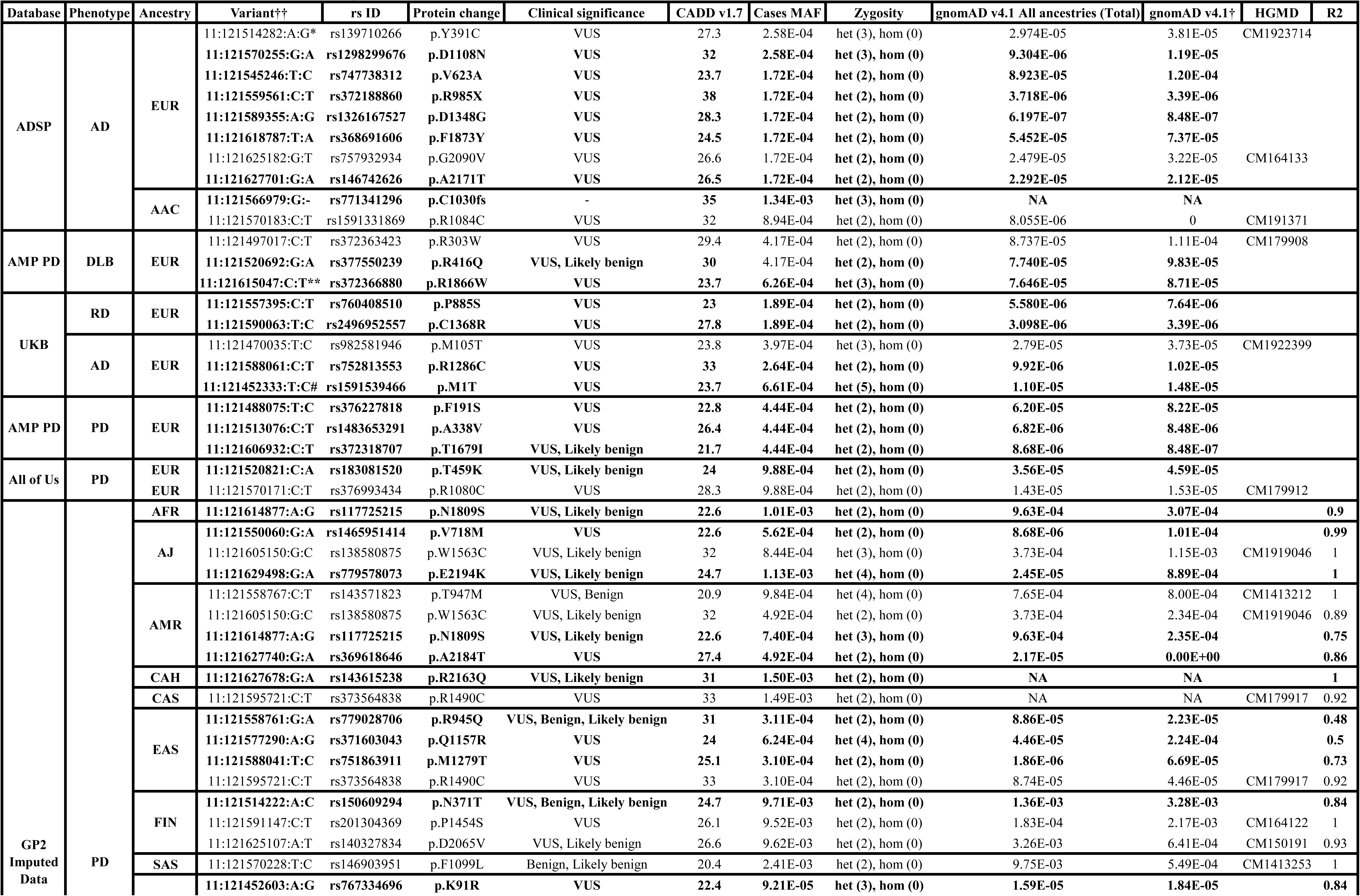
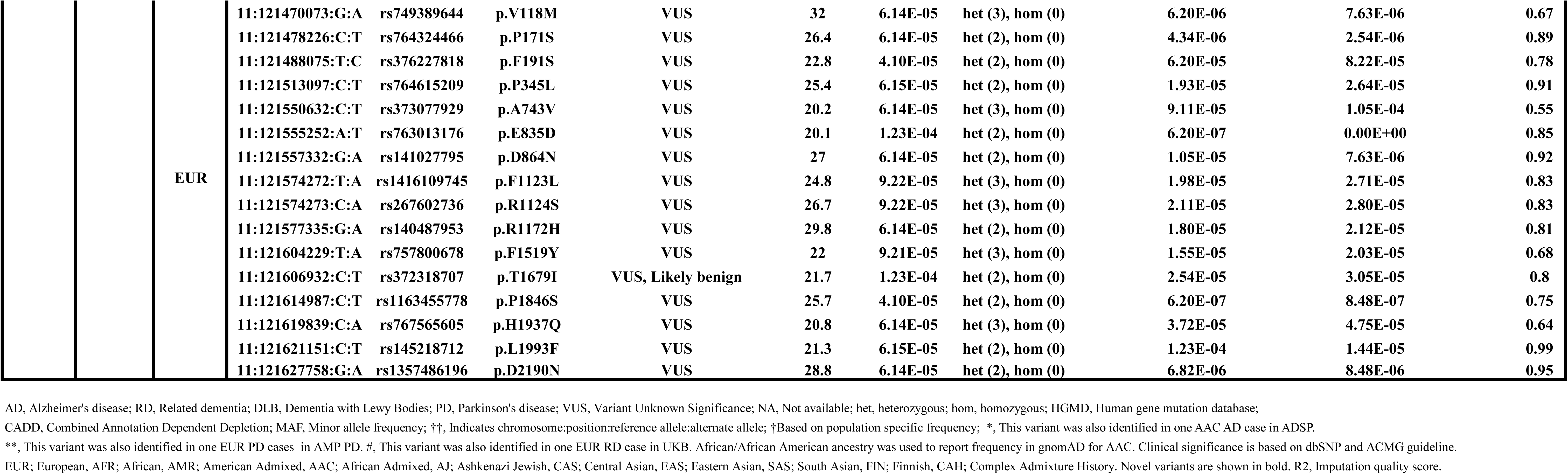
Potentially disease-causing *SORL1* variants present only in Alzheimer’s disease, related dementias, and Parkinson’s disease cases.

Second, we performed association analyses to assess genetic variants that contribute to disease risk. Annotation and functional categorization of the data identified 388 exonic and one splicing variant in ADSP for association analysis. Case-control association analysis nominated 10 variants (two novel and eight previously reported) that reached nominal significance **(Table 2)**. However, none of these associations met the Bonferroni-corrected significance threshold (α = 0.05 / number of variants). Of these, three variants (11:121522975:G:A[p.A528T], 11:121618854:G:A[p.K1895K], 11:121621073:A:G[p.V1967I]) were replicated in AD cases across different ancestries. Gene-based burden analysis did not reveal any significant cumulative effect of *SORL1* variants in populations with sufficient sample sizes (**Supplementary Table 3)**.

**Table 2.** *SORL1* exonic variants with nominal significance in the case-control association analysis of the Alzheimer’s disease, and Parkinson’s disease cohorts.

| Database | Cohort | Variant | rsID | Protein change | Ancestry | A1 | P | OR (L95-U95) | Bonferroni Threshold | gnomAD v4.1 All ancestries (Total) | gnomAD v4.1† | F_A | F_U | R2 |
| --- | --- | --- | --- | --- | --- | --- | --- | --- | --- | --- | --- | --- | --- | --- |
| ADSP | AD | 11:121496917:T:C | rs12364988 | p.H269H | SAS | C | 0.01 | 1.36 (1.07–1.72) | 6.25E-04 | 5.00E-01 | 4.53E-01 | 4.40E-01 | 3.93E-01 |  |
|  |  | 11:121496918:G:A | rs117260922 | p.E270K | CAS†† | A | 0.02 | 117.24 (1.90–7227.41)* | 2.27E-03 | 1.76E-02 | NP | 2.50E-02 | 3.88E-03 |  |
|  |  | 11:121522975:G:A | rs2298813 | p.A528T | AJ | A | 0.04 | 1.70 (1.01–2.85) | 1.85E-03 | 5.70E-02 | 3.21E-02 | 4.00E-02 | 2.51E-02 |  |
|  |  |  |  |  | AAC | A | 0.03 | 1.24 (1.03–1.49) | 3.97E-04 | 5.70E-02 | 9.59E-02 | 1.02E-01 | 8.41E-02 |  |
|  |  | **11:121559627:A:G** | **rs773390553** | **p.M1007V** | **SAS** | **G** | **0.048** | **18.38 (1.03–329.46)\*** | **6.25E-04** | **6.20E-07** | **0** | **2.84E-03** | **2.19E-04** |  |
|  |  | 11:121590137:C:T | rs2276412 | p.N1392N | EUR | T | 0.01 | 0.75 (0.61–0.92) | 2.38E-04 | 2.73E-02 | 1.71E-02 | 1.64E-02 | 2.34E-02 |  |
|  |  | 11:121605213:T:A | rs3824968 | p.A1584A | CAS†† | A | 0.04 | 0.41 (0.17–0.98) | 2.27E-03 | 3.32E-01 | NP | 3.75E-01 | 5.04E-01 |  |
|  |  | **11:121614877:A:G** | **rs117725215** | **p.N1809S** | **EUR** | **G** | **0.02** | **5.98 (1.39–25.85)\*** | **2.38E-04** | **9.63E-04** | **1.11E-03** | **1.72E-03** | **2.27E-04** |  |
|  |  | 11:121614890:T:A | rs62622819 | p.H1813Q | SAS | A | 0.046 | 13.46 (1.05–171.91)* | 6.25E-04 | 8.07E-03 | 6.71E-04 | 2.84E-03 | 4.39E-04 |  |
|  |  | 11:121618854:G:A | rs17125548 | p.K1895K | EUR | A | 0.03 | 0.80 (0.66–0.97) | 2.38E-04 | 3.43E-02 | 2.32E-02 | 2.01E-02 | 2.38E-02 |  |
|  |  |  |  |  | MDE†† | A | 0.04 | 20.80 (1.09–395.99)* | 2.94E-03 | 3.43E-02 | 5.25E-02 | 1.30E-01 | 0 |  |
|  |  |  |  |  | CAH | A | 0.04 | 0.82 (0.67–0.99) | 4.72E-04 | 3.43E-02 | NP | 9.12E-02 | 1.12E-01 |  |
|  |  | 11:121621073:A:G | rs1792120 | p.V1967I | AFR | G | 0.02 | 1.36 (1.05–1.77) | 5.10E-04 | 2.90E-03 | 5.39E-02 | 7.39E-02 | 6.05E-02 |  |
|  |  |  |  |  | AMR | G | 0.03 | 1.96 (1.07–3.60) | 3.36E-04 | 2.90E-03 | 4.00E-03 | 8.29E-03 | 3.78E-03 |  |
| AMP PD | PD | 11:121496918:G:A | rs117260922 | p.E270K | AJ | A | 0.04 | 0.31 (0.10–0.95) | 2.20E-03 | 1.76E-02 | 4.26E-02 | 2.41E-02 | 4.32E-02 |  |
| GP2 Imputed Data | PD | **11:121614877:A:G** | **rs117725215** | **p.N1809S** | **AFR** | **G** | **0.02** | **39.54 (1.82–861.39)\*** | **2.00E-03** | **9.63E-04** | **3.07E-04** | **1.01E-03** | **0** | **0.9** |
|  |  | 11:121522975:G:A | rs2298813 | p.A528T | EAS | A | 0.02 | 0.83 (0.70–0.97) | 1.85E-03 | 5.70E-02 | 1.38E-01 | 1.18E-01 | 1.08E-01 | 1 |
|  |  | **11:121513076:C:T** | **rs1483653291** | **p.A338V** | **EUR** | **T** | **0.047** | **0.16 (0.03–0.97)** | **5.15E-04** | **6.82E-06** | **8.48E-06** | **4.10E-05** | **1.46E-04** | **0.59** |
|  |  | **11:121611093:A:G** | **rs767322041** | **p.I1753V** | **EUR** | **G** | **0.04** | **0.09 (0.00–0.95)** | **5.15E-04** | **8.87E-05** | **1.17E-04** | **3.08E-05** | **2.20E-04** | **0.87** |

\*, The odds ratio may be inflated due to the absence of the variant in controls, low allele count, or limited sample size in this ancestry. Novel variants are shown in bold. †Based on population specific frequency; †† Sample size in cases is less than 50.
AD, Alzheimer's disease; PD, Parkinson's disease; F\_A, Frequency of the affected allele; F\_U, Frequency of the unaffected allele. NP, Not present; African/African American ancestry was used to report frequency in gnomAD for AAC.
EUR; European, AFR; African, AMR; American Admixed, AAC; African Admixed, AJ; Ashkenazi Jewish, CAS; Central Asian, EAS; Eastern Asian, SAS; South Asian, MDE; Middle Eastern, CAH; Complex Admixture History.
The Bonferroni threshold was calculated as 0.05 divided by the number of variants. R2, Imputation quality score.

The AoU analysis included 539 AD cases, 1,655 RD cases and 13,835 controls (**Figure 1**). Although 201 *SORL1* variants were identified in both AD and RD cases, no potential disease-causing variants remained after applying filtering and prioritization criteria in this database (**Table 1, Supplementary Table 1)**.

We further leveraged data from the AMP PD dataset, including 2,522 patients with Dementia with Lewy bodies (DLB) and 3,086 controls (**Figure 1**). Following variant filtering, three potential disease-causing variants in *SORL1* were observed in EUR ancestry individuals: one known variant (11:121497017:C:T[p.R303W]) [36], and two novel variants (11:121520692:G:A[p.R416Q] and 11:121615047:C:T[p.R1866W]) (**Table 1**). No family history was reported for the carriers, and all variants were found in individuals presenting with late onset DLB (AAO ≥65 years) **(Supplementary Table 2)**. Interestingly, the 11:121615047:C:T[p.R1866W] variant was also identified in one male PD patient, with an AAO in the 26-30 year range, no family history, and an Mini-Mental State Examination (MMSE) score of 28, indicating normal cognitive function **(Supplementary Tables 2and 4)**.

The UKB analysis included 3,938 AD cases, 5,586 RD, and 56,684 controls. After filtering, one previously reported variant (11:121470035:T:C[p.M105T]) [37] and four novel *SORL1* variants (11:121557395:C:T[p.P885S], 11:121590063:T:C[p.C1368R], 11:121588061:C:T[p.R1286C], 11:121452333:T:C[p.M1T]) were considered potentially disease causing (**Table 1)**. All carriers were reported to have a late disease onset (≥65 years) **(Supplementary Table 2)**. Among the 14 carriers, 7 (50%) had a family history of AD or PD, while the remaining 7 reported no family history. The variant 11:121452333:T:C[p.M1T], detected in five AD patients, was also observed in one RD case. Interestingly, the known variant 11:121470035:T:C[p.M105T] was found in three LOAD patients: one with a family history of PD, one with a family history of AD, and one without any family history - highlighting the potential contribution of this variant to both AD and PD **(Supplementary Table 2)**.

Finally, WGS data were analyzed from 180 unrelated early-onset dementia cases (including FTD and prion disease) and 3,479 unrelated controls in the 100KGP dataset. Only 11 variants were identified in ADRD cases, and no potential disease-causing variants remained after applying our filtering and prioritization criteria in this database (**Table 1, Supplementary Table 1)**.

All identified variants with CADD > 20, mapped onto the domains and predicted protein structure, are shown in **Figure 2**.

**Figure 2.**
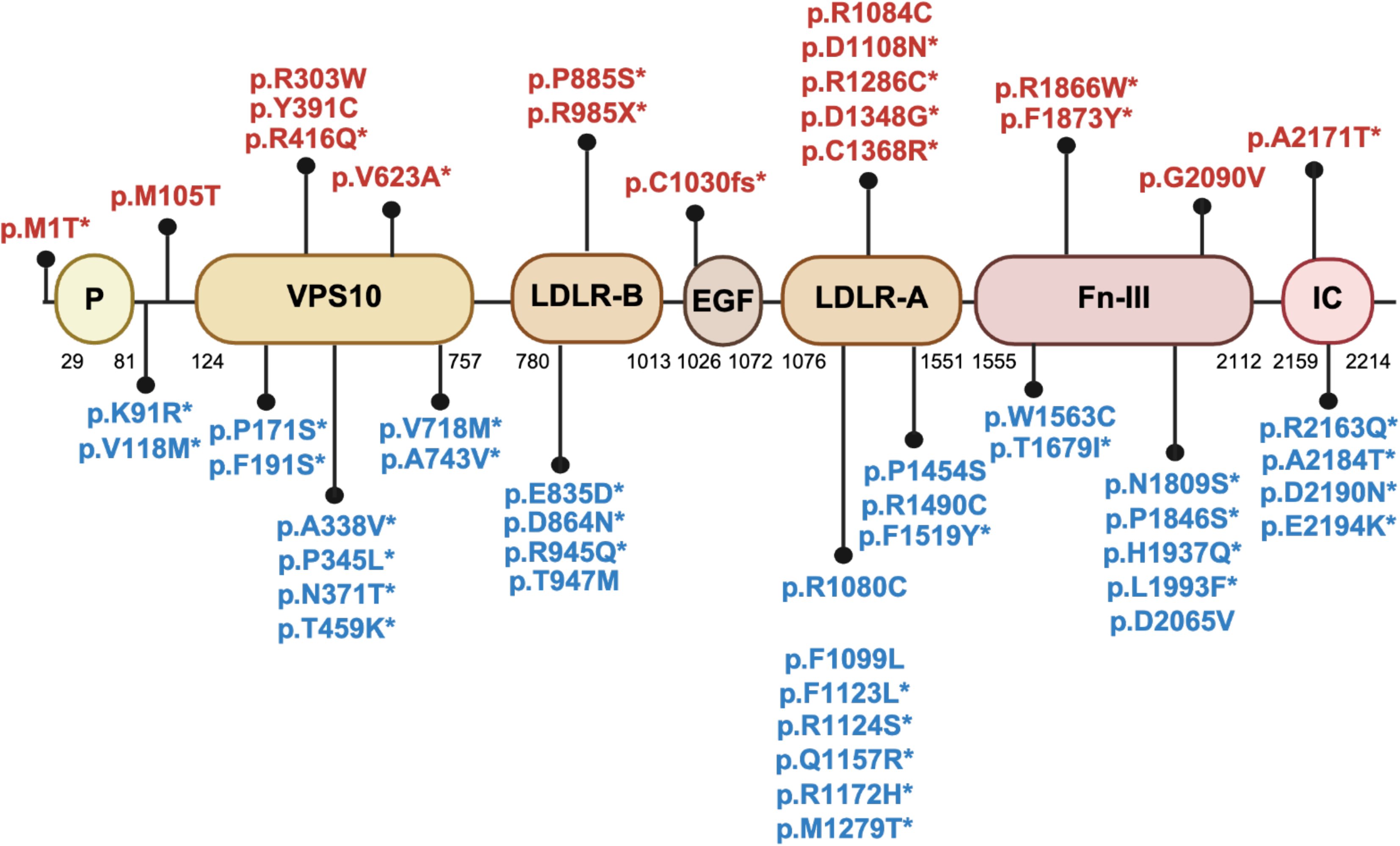
Mutation sites from identified genetic variants with CADD> 20 mapped onto the (A) domains and (B) predicted protein structure encoded by *SORL1*. (A) Graphical representation of *SORL1* variants identified in this study. Variants identified in AD/ADRD are shown at the top, and variants identified in PD are shown at the bottom. Novel variants are marked with an asterisk. EGF: epidermal growth factor like domain; Fn-III: fibronectin-type III domain; IC: intracellular domain; LDLR-A and LDLR-B: low density lipoprotein receptor A and B domains; P: pro-peptide; VPS10: vacuolar protein sorting 10 domain. (B)The predicted protein structure encoded by *SORL1* was obtained from the EMBL AlphaFold Protein Structure Database to ensure that all of the residues were present in protein structure. PyMOL v. 2.6.0 was used to represent the protein structure and its associated mutation sites from identified genetic variants. The yellow color shows beta sheets, the red color shows alpha helices, and the green color shows connecting loops and turns.

### SORL1 variants in Parkinson’s disease

We performed a genetic characterization using a total of 35,233 PD cases and 18,574 controls from 11 ancestries in the GP2 genotyping imputed dataset **(Figure 1)**. Functional annotations of all identified *SORL1* variants by cohort and ancestry in PD before filtering are reported in **Supplementary Table 1**. Following variant filtering, we identified 32 potential disease-causing variants across ancestries (15 with imputation quality (R^2^) ≥ 0.9; **Table 1)**. Among these, five had been previously reported in AD: 11:121605150:G:C[p.W1563C], 11:121558767:C:T[p.T947M], 11:121595721:C:T[p.R1490C], 11:121591147:C:T[p.P1454S], and 11:121570228:T:C[p.F1099L] and, one in PD: 11:121625107:A:T[p.D2065V] [11,36,38–40]. The remaining 26 variants were novel. Details of clinical data for all carriers are reported in **Supplementary Table 2 and 4**. Among the 86 carriers, 25 (29.1%) reported a family history of PD, 45 (52.3%) reported no family history, and family history data were unavailable for 16 (18.6%). AAO data were available for 52 carriers, of whom 35 (67.3%) were classified as late-onset PD (AAO ≥50 years) and 17 (32.7%) as early-onset PD (AAO <50 years), while AAO data were missing for 34 individuals. Among carriers with available cognitive data, most did not show cognitive impairment, except for three individuals: one carrier of 11:121470073:G:A[p.V118M] had a Montreal Cognitive Assessment (MoCA) score of 24, consistent with PD-MCI; one carrier of 11:121606932:C:T[p.T1679I] had an MMSE score of 20, indicating moderate cognitive impairment; and one carrier of 11:121605150:G:C[p.W1563C] had a MoCA score of 22, suggestive of PD-MCI.

Annotation and functional categorization identified 145 unique exonic variants and one splicing variant in the dataset for association analysis, which revealed four variants (three novel and one previously reported) that reached nominal significance across all ancestries **(Table 2)**. However, none of these associations met the Bonferroni-corrected significance threshold (α = 0.05 / number of variants) **(Table 2)**. Gene-level burden analysis indicated no significant cumulative effect in PD (**Supplementary Table 3**).

We further analyzed sequencing data from the AMP PD WGS cohort, which included 2,600 PD cases and 3,180 controls. After filtering, three novel potential disease-causing variants remained (11:121488075:T:C[p.F191S], 11:121513076:C:T[p.A338V], 11:121606932:C:T[p.T1679I]) **(Table 1, Supplementary Table 1)**. Among the six carriers, three reported no family history of PD, one reported a family history of PD, and family history data were unavailable for two individuals. All six identified individuals were diagnosed with classical PD. The MMSE and MoCA assessments showed scores within the normal range across multiple visits for all individuals. Only one carrier of 11:121488075:T:C[p.F191S] showed a slight decline in MoCA scores—from 29 to 26 over three years—which, while still within the normal range, may indicate subtle cognitive changes that may warrant further follow up. Another individual, carrier of 11:121513076:C:T[p.A338V], self-reported slight cognitive impairment; however, this did not impact the MoCA score. More detailed clinical data on motor and non-motor signs are reported in **Supplementary Table 4**. Overall, the clinical data support a diagnosis of typical PD without evidence of relevant cognitive impairment.

Annotation and functional categorization of the data identified 112 unique exonic variants and no splicing variants in AMP PD for association analysis. Association analysis was performed in both EUR and AJ ancestries. In the AJ ancestry, one variant (11:121496918:G:A[p.E270K]) showed a nominally significant association (odds ratio (OR)=0.31, 95% CI=0.1-0.95, P=0.04), suggesting a potential protective effect **(Table 2)**. However, this association did not meet the Bonferroni-corrected significance threshold of P < 0.0022 (α = 0.05 / number of variants). No significant associations were observed in EUR ancestry. Gene-based burden analysis did not reveal any significant cumulative effects of the variants in *SORL1* in PD (**Supplementary Table 3)**.

The All of Us analysis included 1,422 PD cases and 13,835 controls **(Table 1)**. After filtering, one previously reported variant (11:121570171:C:T[p.R1080C]) [36] and one novel variant (11:121520821:C:A[p.T459K]) remained as potential disease causing **(Table 1, Supplementary Table 1)**. Both variants were identified in two PD patients each. For both variants, one patient had no family history of PD, and family history information was unavailable for the other. Based on the available clinical data, they did not have a diagnosis of cognitive impairment or dementia; however, detailed data and cognition rating scale scores were unavailable.

The UKB analysis included 2,938 PD cases, and 56,684 controls (**Figure 1**). Although many *SORL1* variants were identified in PD cases, no potentially disease-causing variants remained following our filtering criteria in this database **(Table 1, Supplementary Table 1)**.

WGS data in 100KGP from 570 unrelated PD cases, and 3,479 unrelated controls were analyzed. We identified 14 variants in *SORL1,* none of which remained as potentially disease-causing after applying our filtering criteria **(Table 1, Supplementary Table 1)**.

All identified variants with CADD > 20, mapped onto the domains and predicted protein structure, are shown in **Figure 2**.

### Discovery of SORL1 variants in Parkinson’s disease across global populations suggests shared genetic susceptibility with Alzheimer’s disease

We conducted a multi-ancestry, comprehensive genetic characterization of *SORL1,* which identified several novel and previously reported variants across different datasets and populations. Two novel variants, 11:121488075:T:C[p.F191S] and 11:121606932:C:T[p.T1679I], were detected in EUR individuals with PD in both the AMP PD and GP2 genotyped imputed datasets. Another novel variant, 11:121614877:A:G[p.N1809S], was identified in the GP2 genotyped imputed data among the AFR and AMR populations. Notably, this variant was also associated with AD in EUR individuals in the ADSP dataset. Among the previously reported variants, 11:121605150:G:C[p.W1563C] was initially identified in an AJ family diagnosed with AD [38]; in our analysis, we found this variant in PD patients within the GP2 genotyped imputed dataset, specifically among AJ and AMR populations. Similarly, 11:121595721:C:T[p.R1490C], previously reported in Dutch AD cohorts [36], was identified in PD patients from CAS and EAS populations within the GP2 dataset. Replication of the novel variants in individuals with PD and across different ancestries supports their potential role in disease causation. Furthermore, the replication of variants in both PD and AD suggests possible shared genetic risk etiologies and highlights their broader relevance for therapeutic development.

Two variants previously reported in AD cases from EUR populations were also detected in AD cases of the same ancestry group. These variants include 11:121514282:A:G[p.Y391C] [34] and 11:121625182:G:T[p.G2090V] [11]. In addition, 11:121470035:T:C[p.M105T], which was reported in an AD case from a Northern European ancestry group with a strong family history of AD, was also identified in our study in EUR AD cases, including one individual with a family history of PD [37]. Variant 11:121591147:C:T[p.P1454S] and 11:121570228:T:C[p.F1099L], previously reported in AD cases of EUR ancestry [11,39], were identified in our study in PD cases of FIN and SAS ancestries, respectively. Variants 11:121497017:C:T[p.R303W] and 11:121570171:C:T[p.R1080C], both previously reported in Dutch AD cases [36], were identified in our study in individuals with DLB and PD, respectively, within EUR ancestry. Variant 11:121570183:C:T[p.R1084C], reported in AD cases from the Saudi population [35], was identified in our study in AD cases of AAC ancestry; 11:121558767:C:T[p.T947M], previously reported in AD among Caribbean Hispanics [39], was observed in PD cases of AMR ancestry; and 11:121625107:A:T[p.D2065V], found in PD cases of EUR populations [40], was identified in PD cases of FIN ancestry. The latter identification confirms the role of the 11:121625107:A:T[p.D2065V] variant in PD. The frequency of all of these variants in our ancestry-based cohorts was higher than the ancestry-specific frequencies reported in gnomAD v4.1.0.

Association analysis identified several variants that were replicated across cohorts and databases. For example, 11:121496918:G:A[p.E270K], a previously reported variant in AD [39], was found in PD cases of AJ ancestry in the AMP PD dataset. Another previously reported variant in AD (11:121522975:G:A[p.A528T]), which was found in the Caribbean Hispanic population [39], was replicated in the AD cohorts of the AJ and AAC ancestries and in the PD cohort of the EAS ancestry. Interestingly, both variants were associated with a reduced risk of PD, and these results should be interpreted with caution (**Table 2**).

Interestingly, we identified one previously reported variant — 11:121522975:G:A[p.A528T] (OR = 0.83, 95% CI=0.7–0.97, P=0.02) — and two novel variants — 11:121611093:A:G[p.I1753V] (OR=0.09, 95% CI=0.00-0.95, P=0.04), and 11:121513076:C:T[p.A338V] (OR=0.16, 95% CI=0.03-0.97, P=0.047) — associated with reduced risk of PD in EAS, EUR, and EUR populations, respectively. Of note, 11:121522975:G:A[p.A528T] was found to increase the risk of AD in the AJ (OR=1.7, 95% CI=1.01-2.85, P=0.04) and AAC (OR=1.24, 95% CI=1.03-1.49, P=0.03) populations, suggesting it may have population-specific effects; however, the nominal significance indicates that these associations should be interpreted with caution and require replication (**Table 2**).

### Risk-reducing variants for Alzheimer’s disease identified in multiple populations

Two variants, 11:121590137:C:T[p.N1392N] and 11:121618854:G:A[p.K1895K], have previously been associated with decreased LOAD risk in European populations [41]. In our study, we identified 11:121590137:C:T[p.N1392N] with an OR of 0.75 (95% CI=0.61-0.92, P=0.01) in the EUR population, and 11:121618854:G:A[p.K1895K] with an OR of 0.80 (95% CI=0.66-0.97, P=0.03) in EUR and an OR of 0.82 (95% CI=0.67-0.99, P=0.04) in the CAH population. While some overlap may exist between our European dataset and that of Liu et al., these results are generally consistent with previous findings and, tentatively, extend the association to the CAH ancestry. However, because CAH represents a highly admixed population with potential case–control substructure imbalance, this observation should be interpreted with caution, as it may partly reflect population heterogeneity rather than a true ancestry-specific effect. Taken together, these findings support a protective effect of the T and A alleles, respectively, against AD risk **(Table 2)**.

### Family-based analysis of SORL1 identified rare predicted-damaging variants in Parkinson’s disease

We used 3,998 WGS-genotyped individuals from the GP2 release 10 monogenic WGS data and reconstructed a total of 469 families, which are composed of 335 core pedigrees (defined by the Monogenic Working Group) and 134 additional kinship-inferred families with at 2 or more family members. We next merged these two family groups into a unified pedigree set of 398 informative families (900 samples) that had at least one PD case and at least two total members. Screening the informative WGS pedigree set identified 72 individuals across 40 families carrying an established PD-associated variant and/or a ClinVar pathogenic/likely-pathogenic variant in a known PD gene. Out of the 900 samples, only one had a diagnosis of PDD. We observed the largest numbers of families in EUR ancestry (594 samples) and EAS (159 samples), and smaller numbers from CAS, SAS, AMR, AFR, CAH, FIN and MDE. Next, we obtained 362 distinct rare, predicted-damaging *SORL1* variants (LOF, missense with CADD > 20, with gnomAD v4.1 allele frequency < 1%, **Supplementary Table 5**) from the WGS data of the informative families. We selected *SORL1* variants that showed co-segregation with PD (**Table 3, Supplementary Figure 1**). In EAS individuals, variant 11:121478242:G:A [p.R176Q] was observed in two independent families, one with a pair of affected siblings and another with a parent-child duo. In the family with the pair of affected siblings, both carry the *GBA1* RecNcil allele, which includes the pathogenic p.L483P variant in heterozygous state. In EUR individuals, variant 11:121514222:A:C [p.N371T] was detected in a single family with three affected siblings, all carriers. None of these three EAS and EUR family members carry any known PD variant or pathogenic variant in a known PD gene. One additional variant (11:121545392:G:A, p.V672M) was present in a single family of EUR ancestry with one affected pair of siblings, and complete carriership among them. This family also carries the known PD variant p.G192R in the *RAB39B* gene [42]. None of these selected families have members diagnosed with dementia. We further evaluated the association of these variants in unrelated case-control samples from the WGS dataset including 10,261 individuals of which EAS: 1,793 PD and 346 controls; EUR: 6,782 PD and 1,340 controls. Although none showed statistically significant enrichment in PD (**Supplementary Table 6**), all three variants had slightly higher carrier frequencies in cases than controls: p.R176Q (EAS; 4/1,793 [2.23E-3] vs. 0/346 [0]), p.N371T (EUR; 25/6,781 [3.69E-3] vs. 3/1,340 [2.24E-3]), and p.V672M (EUR; 2/6,782 [2.95E-4] vs. 0/1,339 [0]). In addition to the two EAS families carrying p.R176Q, the unrelated case-control cohort added two additional PD carriers (including one familial PD singleton), and both variants identified in EUR individuals (p.N371T and p.V672M) also had carriers outside the informative pedigrees. Together, these findings show a set of rare, ancestry-specific *SORL1* variants segregating with PD, with a tendency for higher frequency in unrelated PD cases vs. controls.

**Table 3.** Rare, predicted-damaging SORL1 variants that co-segregated with Parkinson’s disease in the family-based analysis.

| Ancestry | Variant_ID | Protein change | CADD v1.7 | Families_with_carriers | Zygosity | Family_members | PD_cases_carriers | gnomAD_AF_ancestry | CC_PD_carriers/PD_total | CC_ctrl_carriers/ctrl_total |
| --- | --- | --- | --- | --- | --- | --- | --- | --- | --- | --- |
| EAS | 11:121478242:G:A | p.R176Q | 28.6 | 2 | Het | 4 | 4 | 5.00E-04 | 4/1793 (2.23E-3) | 0/346 (0) |
| EUR | 11:121514222:A:C | p.N371T | 24.7 | 1 | Het | 3 | 3 | 2.50E-03 | 25/6781 (3.69E-3) | 3/1340 (2.24E-3) |
| EUR | 11:121545392:G:A | p.V672M | 23.5 | 1 | Het | 2 | 2 | 1.35E-05 | 2/6782 (2.95E-4) | 0/1339 (0) |

Variant\_ID: GRCh38 variant identifier (chromosome, base pair, reference allele, alternate allele). Families\_with\_carriers: Number of pedigrees with at least one PD carrier of the variant. Family\_members: Total number of family members in those pedigrees. PD\_cases\_carriers: Number of PD cases carrying the variant in those pedigrees.
gnomAD\_AF\_ancestry: gnomAD v4.1 ancestry-specific allele frequency for the variant. CC\_PD\_carriers/PD\_total: Number and proportion (in parentheses) of carriers among PD cases in the unrelated GP2 Release 10 case–control cohort.
CC\_ctrl\_carriers/ctrl\_total: Number and proportion (in parentheses) of carriers among controls in the unrelated GP2 Release 10 case–control cohort. EAS, East Asian; EUR, European, Het, Heterozygous.

### Fine-mapping supports rs11218343 as the lead SORL1 variant in Europeans

Fine-mapping identified only one SNP, rs11218343, with a PP of 0.99 and a p-value of 1.01 × 10^-14^ in individuals of EUR ancestry from the Bellenguez et al. GWAS dataset. In the Kunkle et al., 2021 [29], meta-analyzed PD data, MVP AD, and MVP ADRD-AD-PD GWAS summary statistics, fine-mapping analysis favored the null model (no association in the region) (PP > 0.6), suggesting that none of the variants tested in these regions are likely to be causally associated.

### No significant effects of identified variants on SORL1 or nearby gene expression in brain tissue

Functional annotation of the variants identified in the association analysis, using multivariate brain eQTL data, did not reveal any significant associations (p < 5×10^-8^) and provided no evidence that these variants affect the expression of *SORL1* or nearby genes **(Supplementary Table 7)**.

## Discussion

The *SORL1* gene has been widely recognized as a prominent target for therapeutic development in AD. As we enter a new era of precision therapeutics, true precision remains elusive without a comprehensive, global understanding of disease. Neurodegeneration represents a spectrum of disorders with overlapping features, making it essential to conduct research across the full range of conditions within this umbrella to uncover shared mechanisms and improve diagnosis and treatment. Notably, two-thirds of FDA-approved drugs in 2021 are supported by human genetic evidence, highlighting the importance of genetic insights in drug development [43].

Here, we conducted a large-scale, comprehensive analysis of potential disease-causing *SORL1* variants associated with ADRDs and PD, leveraging the largest multi-ancestry, biobank-scale dataset to date. We identified 53 potential disease-causing *SORL1* variants across all cohorts, including 41 novel and 12 previously reported. Of these, 18 variants were detected in ADRD cases (13 novel, 5 previously reported), and 35 in PD cases (28 novel, 7 previously reported).

Five variants present in PD cases (11:121488075:T:C[p.F191S], 11:121606932:C:T[p.T1679I], 11:121614877:A:G[p.N1809S], 11:121605150:G:C[p.W1563C], 11:121595721:C:T[p.R1490C]) were replicated across multiple ancestries and databases, suggesting a role of this gene in PD. The role of *SORL1* in DLB remains unclear. A previous study [44] reported rare, damaging *SORL1* variants in DLB but did not observe a statistically significant enrichment, largely because these variants were singletons or carried by very few individuals. In our analysis, we identified three *SORL1* variants (11:121497017:C:T[p.R303W], 11:121520692:G:A[p.R416Q], and 11:121615047:C:T[p.R1866W]) in individuals diagnosed with DLB, observed in two, two, and three carriers, respectively; one of these variants was also seen in a PD case without dementia. These same variants were also reported by Ray et al [44]. Although the evidence remains limited due to low carrier counts, the recurrence of these rare variants across studies supports continued investigation into the potential contribution of *SORL1* to DLB and related neurodegenerative phenotypes. One limitation of this study is that individuals were not systematically screened for pathogenic variants in other known AD/PD genes and may therefore harbor variants in additional genes that contribute to disease risk. Association analyses identified 10 nominally significant variants for AD (2 novel, 8 known) (11:121496917:T:C[p.H269H], 11:121496918:G:A[p.E270K], 11:121522975:G:A[p.A528T], 11:121559627:A:G[p.M1007V], 11:121590137:C:T[p.N1392N], 11:121605213:T:A[p.A1584A], 11:121614877:A:G[p.N1809S], 11:121614890:T:A[p.H1813Q], 11:121618854:G:A[p.K1895K], 11:121621073:A:G[p.V1967I]) and five for PD (3 novel, 2 known) (11:121496918:G:A[p.E270K], 11:121614877:A:G[p.N1809S], 11:121522975:G:A[p.A528T], 11:121611093:A:G[p.I1753V], 11:121513076:C:T[p.A338V]), with three variants showing nominal significance in both AD and PD cohorts (11:121496918:G:A[p.E270K], 11:121522975:G:A[p.A528T],11:121614877:A:G[p.N1809S]).

Regarding the variants associated with both AD and PD identified in this study, the collected evidence indicates that the associations for certain variants are more robust than for others. These include 11:121522975:G:A[p.A528T] in AD within the AAC ancestry, 11:121618854:G:A[p.K1895K] in AD in CAH ancestry, and 11:121621073:A:G[p.V1967I] in AD among individuals of AFR and AMR ancestries. For PD, stronger associations were observed for 11:121496918:G:A[p.E270K] in AJ ancestry, and 11:121522975:G:A[p.A528T] in EAS. The previous reports of these variants reinforce the reliability of our findings across different populations.

Among the identified variants, some warrant cautious interpretation due to limited supporting data. For instance, while the novel variant 11:121513076:C:T[p.A338V] appears to be associated with PD in individuals of EUR ancestry, its low imputation quality (R^2^=0.59) raises concerns about reliability. Similarly, the novel variant 11:121614877:A:G[p.N1809S], identified in individuals of AFR ancestry from the GP2 dataset (995 cases, 1,671 controls), showed a significant association (P = 0.02, OR = 39.54; F-A (frequency in affected individuals) = 1.01 × 10^-3^, F-U (frequency in unaffected individuals) = 0). Although the imputation quality for this variant is high (R^2^=0.90), the result is based on imputed data, which are inherently probabilistic and should therefore be interpreted with caution. The same variant was also observed in individuals of EUR ancestry in the AD cohort (5,812 cases and 4,411 controls), where it showed a rare allele frequency (F-A = 1.72 × 10^-3^, F-U = 2.27 × 10^-4^) and was associated with an OR of 5.98 (95% CI: 1.39–25.85, p = 0.02). Despite the relatively large sample size, the low allele count suggests that this estimate should also be interpreted cautiously and validated in independent cohorts. Interpretation is limited by small sample sizes and wide confidence intervals for 11:121496918:G:A[p.E270K] (CAS), 11:121559627:A:G[p.M1007V] (SAS), 11:121614890:T:A[p.H1813Q] (SAS), and 11:121618854:G:A[p.K1895K] (MDE). Another important consideration is the frequency of identified variants in cases relative to the general population, such as gnomAD, which can influence interpretation. This comparison indicates that several previously reported variants do not show notable enrichment in cases. For example, 11:121496917:T:C[p.H269H] (SAS), 11:121522975:G:A[p.A528T] (AJ), 11:121590137:C:T[p.N1392N] (EUR), 11:121605213:T:A[p.A1584A] (CAS), and 11:121618854:G:A[p.K1895K] (EUR) all had case frequencies comparable to population-specific or general gnomAD frequencies. Similarly, the novel 11:121611093:A:G[p.I1753V] was extremely rare in both cases and gnomAD populations. These observations underscore the importance of considering background allele frequencies when evaluating potential disease associations.

Two *SORL1* variants (11:121496918:G:A[p.E270K], 11:121522975:G:A[p.A528T]), previously associated with AD risk, were identified in PD cases for the first time in our study but with opposite effects. Both were nominally significant and require replication in independent PD cohorts. The nominal nature of these associations, coupled with the opposite effect estimates between AD and PD, suggests that these results should be interpreted with caution. 11:121470035:T:C[p.M105T] was also identified in an AD patient with a family history of PD. These findings suggest that *SORL1* contributes to neurodegeneration more broadly, but with potentially divergent effects on risk across AD and PD.

Despite these findings, no significant cumulative enrichment at a gene level was observed across ancestries with sufficient sample size in either disease group. Notably, the previously reported *SORL1* variant linked to PD in a Greek family was not detected in our dataset.

In the family-based analysis we identified three rare, predicted-damaging *SORL1* variants that co-segregated with PD in four small families across two ancestries. It is important to note that one EAS family with the p.R176Q variant also carry the pathogenic *GBA1* p.L483P variant in their two affected siblings, and that one EUR pedigree, carrying p.V672M, harbored the known PD variant p.G192R in *RAB39B*; thus, the p.R176Q and p.V672M segregation signals should be interpreted cautiously with respect to causality. Only the p.N371T variant has been previously associated with Alzheimer’s disease and Frontotemporal Lobar Degeneration [45,46]. Although these variants segregated within pedigrees and were marginally more frequent in cases than controls, none showed significant enrichment in unrelated case–control cohorts, showing the limited power for associations of ultra-rare variants. This pattern is consistent with prior reports [18], illustrating how rare damaging variants may segregate in families while remaining undetectable in general association tests.

Taken together, our findings support an expanded role for *SORL1* in neurodegeneration beyond AD and reinforce the importance of diverse populations in genetic risk evaluation.

## Abbreviations

100KGP: 100,000 Genomes Project AAC African Admixed
AAO: Age At Onset
AD: Alzheimer’s Disease
ADGC: Alzheimer’s Disease Genetics Consortium
ADSP: Alzheimer’s Disease Sequencing Project
AFR: African
AJ: Ashkenazi Jewish
AMR: American Admixed
AMP PD: Accelerating Medicines Partnership in Parkinson’s Disease
AoU: All of Us
APP: Amyloid Precursor Protein
CADD: Combined Annotation Dependent Depletion
CAH: Complex Admixture History
CAS: Central Asian
DLB: Dementia with Lewy bodies
EAS: Eastern Asian
EOAD: Early-Onset AD
EUR: European
FIN: Finnish
GnomAD: Genome Aggregation Database
GP2: Global Parkinson’s Genetics Program
GWAS: Genome-Wide Association Studies
LOAD: Late-Onset AD
LOF: loss-of-function
MAC: Minor Allele Count
MDE: Middle Eastern
MMSE: Mini-Mental State Examination
MoCA: Montreal Cognitive Assessment
MVP: Million Veteran Program
OR: Odds Ratio
PCs: Principal Components
PD: Parkinson’s Disease
PP: Posterior Probability
RD: Related Dementias
SAS: South Asian
SKAT-O: Optimized nonburden Sequence Kernel Association Test
SORL1: Sortilin-Related Receptor
UKB: United Kingdom Biobank
WGS: Whole-Genome Sequencing

## Acknowledgments

We thank Paige Brown Jarreau for her meticulous editing of this manuscript.

This research was supported [in part] by the Intramural Research Program of the National Institutes of Health (NIH); project number ZO1 AG000535 and ZIA AG000949. The contributions of the NIH author(s) were made as part of their official duties as NIH federal employees, are in compliance with agency policy requirements, and are considered Works of the United States Government. However, the findings and conclusions presented in this paper are those of the author(s) and do not necessarily reflect the views of the NIH or the U.S. Department of Health and Human Services. This work utilized the computational resources of the NIH HPC Biowulf cluster. (http://hpc.nih.gov).

We have received an exception to the Data and Statistics Dissemination Policy from the All of Us Resource Access Board. The All of Us Research Program is supported by the National Institutes of Health, Office of the Director: Regional Medical Centers: 1 OT2 OD026549; 1 OT2 OD026554; 1 OT2 OD026557; 1 OT2 OD026556; 1 OT2 OD026550; 1 OT2 OD 026552; 1 OT2 OD026553; 1 OT2 OD026548; 1 OT2 OD026551; 1 OT2 OD026555; IAA #: AOD 16037; Federally Qualified Health Centers: HHSN 263201600085U; Data and Research Center: 5 U2C OD023196; Biobank: 1 U24 OD023121; The Participant Center: U24 OD023176; Participant Technology Systems Center: 1 U24 OD023163; Communications and Engagement: 3 OT2 OD023205; 3 OT2 OD023206; and Community Partners: 1 OT2 OD025277; 3 OT2 OD025315; 1 OT2 OD025337; 1 OT2 OD025276. In addition, the All of Us Research Program would not be possible without the partnership of its participants.

This research has been conducted using the UK Biobank Resource under application number 33601.

We gratefully acknowledge the participants of the National Genomic Research Library (NGRL), whose contributions made this research possible. Secure access to the NGRL under project ID [RR1206] was provided by Genomics England, which delivers the NGRL in partnership with NHS England, and is wholly owned by the UK Department of Health and Social Care. The NGRL contains participants’ health data collected by the NHS as part of their care, along with samples and data from their participation in research, for which fully informed consent has been obtained. This includes genomic and clinical data provided through the NHS Genomic Medicine Service, as well as data obtained through research studies, including the 100,000 Genomes Project and the Generation Study, both of which are delivered in partnership with the NHS, and from other research cohorts involving external collaborators.

This research has been conducted using the Alzheimer’s Disease Sequencing Project (ADSP) Resource under accession number NG00067. The data for this study were prepared, archived, and distributed by the National Institute on Aging Alzheimer’s Disease Data Storage Site (NIAGADS) at the University of Pennsylvania (U24-AG041689), funded by the National Institute on Aging.

This project was supported by the Global Parkinson’s Genetics Program (GP2; https://gp2.org). GP2 is funded by the Aligning Science Across Parkinson’s (ASAP) initiative and implemented by The Michael J. Fox Foundation for Parkinson’s Research (MJFF). For a complete list of GP2 members see https://doi.org/10.5281/zenodo.7904831.

Data used in the preparation of this article were obtained from the AMP PD Knowledge Platform. For up-to-date information on the study, please visit https://www.amp-pd.org. AMP PD—a public-private partnership—is managed by the FNIH and funded by Celgene, GSK, the Michael J. Fox Foundation for Parkinson’s Research, the National Institute of Neurological Disorders and Stroke, Pfizer, Sanofi, and Verily. Clinical data and biosamples used in the preparation of this article were obtained from the Parkinson’s Progression Markers Initiative (PPMI), and the Parkinson’s Disease Biomarkers Program (PDBP). PPMI—a public-private partnership—is funded by the Michael J. Fox Foundation for Parkinson’s Research and funding partners, including full names of all of the PPMI funding partners found at http://www.ppmi-info.org/fundingpartners. The PPMI Investigators have not participated in reviewing the data analysis or content of the manuscript. For up-to-date information on the study, visit http://www.ppmi-info.org. The Parkinson’s Disease Biomarker Program (PDBP) consortium is supported by the National Institute of Neurological Disorders and Stroke (NINDS) at the National Institutes of Health. A full list of PDBP investigators can be found at https://pdbp.ninds.nih.gov/policy. The PDBP Investigators have not participated in reviewing the data analysis or content of the manuscript. PDBP sample and clinical data collection is supported under grants by NINDS: U01NS082134, U01NS082157, U01NS082151, U01NS082137, U01NS082148, and U01NS082133.

## Author Contributions

SBC, contributed to the study concept or design. MK, SNY, CCC, AK, BIB, SMG, SCA, FA, PRP, and ZS, were involved in the analysis of data across different biobanks. PSL, Protein Structure Modeling. LML, Visualization and Interpretation of clinical data. All authors contributed to the critical review and had final responsibility for the decision to submit for publication.

## Declarations

### Ethics approval and consent to participate

Research conducted on biobank-scale data was deemed ‘not human subjects research’ by the NIH Office of IRB Operations and stated that no IRB approval is required. The NIH Intramural IRB waived ethical approval for the overall study.

### Consent for publication

Not applicable.

### Data availability

All data supporting the findings of this study are available within the paper, its Supplementary Information files, and the repositories listed below.

Data used in the preparation of this article were obtained from the Global Parkinson’s Genetics Program (GP2; https://gp2.org). Specifically, we used Tier 2 data from GP2 release 9 (DOI: 10.5281/zenodo.14510099) and release 10 (DOI: 10.5281/zenodo.15748014). GP2 data can be requested through AMP PD (https://amp-pd.org).

Data were also used from:

- All of Us (AoU) v7 (https://www.researchallofus.org/register/)
- Alzheimer’s Disease Sequencing Project (ADSP) v4 (https://adsp.niagads.org/)
- Accelerating Medicines Partnership – Parkinson’s Disease (AMP PD) v3 (https://amp-pd.org/register-for-amp-pd)
- 100,000 Genomes Project (100KGP) v18 (https://www.genomicsengland.co.uk/)
- UK Biobank (UKB) v18.1 (https://www.ukbiobank.ac.uk/enable-your-research/register)
- Global Parkinson’s Genetics Program (GP2) releases v9 and v10 (https://amp-pdrd.org/register-for-amp-pd)
- Summary statistics from Bellenguez *et al.* (2022), Kunkle *et al.* (2021), Shigemizu *et al.* (2021), Sherva *et al.* (2023), and Leonard *et al.* (2025)
- FinnGen Release 12 (https://www.finngen.fi/en)
- Multi-variate multiple QTL (mmQTL) dataset by Zeng *et al*. (2022)

### Code availability

All code generated for this article, and the identifiers for all software programs and packages used, are available on GitHub (https://github.com/GP2code/SORL1_ADPD) and were given a persistent identifier via Zenodo (DOI 10.5281/zenodo.17819222).

---

**Supplementary Figure 1.**
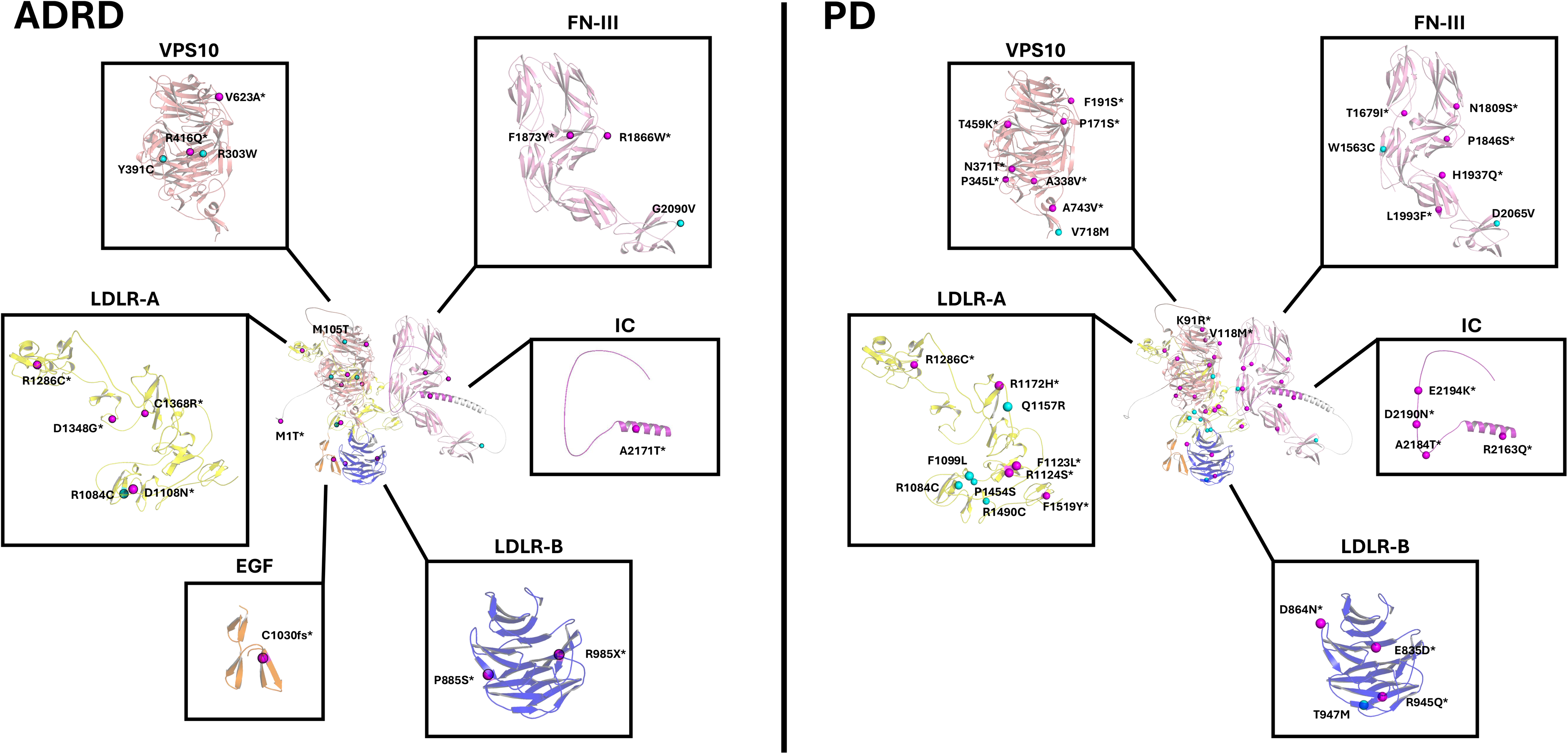
*SORL1* candidate-variant carrier pedigrees. (A) Two East Asian (EAS) families carrying *SORL1* p.R176Q (11:121478242:G:A). (B) European (EUR) family carrying *SORL1* p.N371T (11:121514222:A:C). (C) European (EUR) family carrying *SORL1* p.V672M (11:121545392:G:A). Squares indicate males and circles indicate females; filled symbols denote individuals affected with PD, unfilled symbols indicate samples without genetic data available. Age at onset (AAO) is shown when available. Genotype is shown as Wt/Mut (heterozygous) for the respective *SORL1* variant. ^#^ indicates individual carriers of the PD associated mutation p.L483P in *GBA1*. * indicates individual carriers of the PD associated mutation p.G192R in *RAB39B*.

