## Supplementary figures and images for "Is *SORL1* a common genetic target across neurodegenerative diseases?: A multi-ancestry biobank scale assessment"

### Supplementary Figure 1

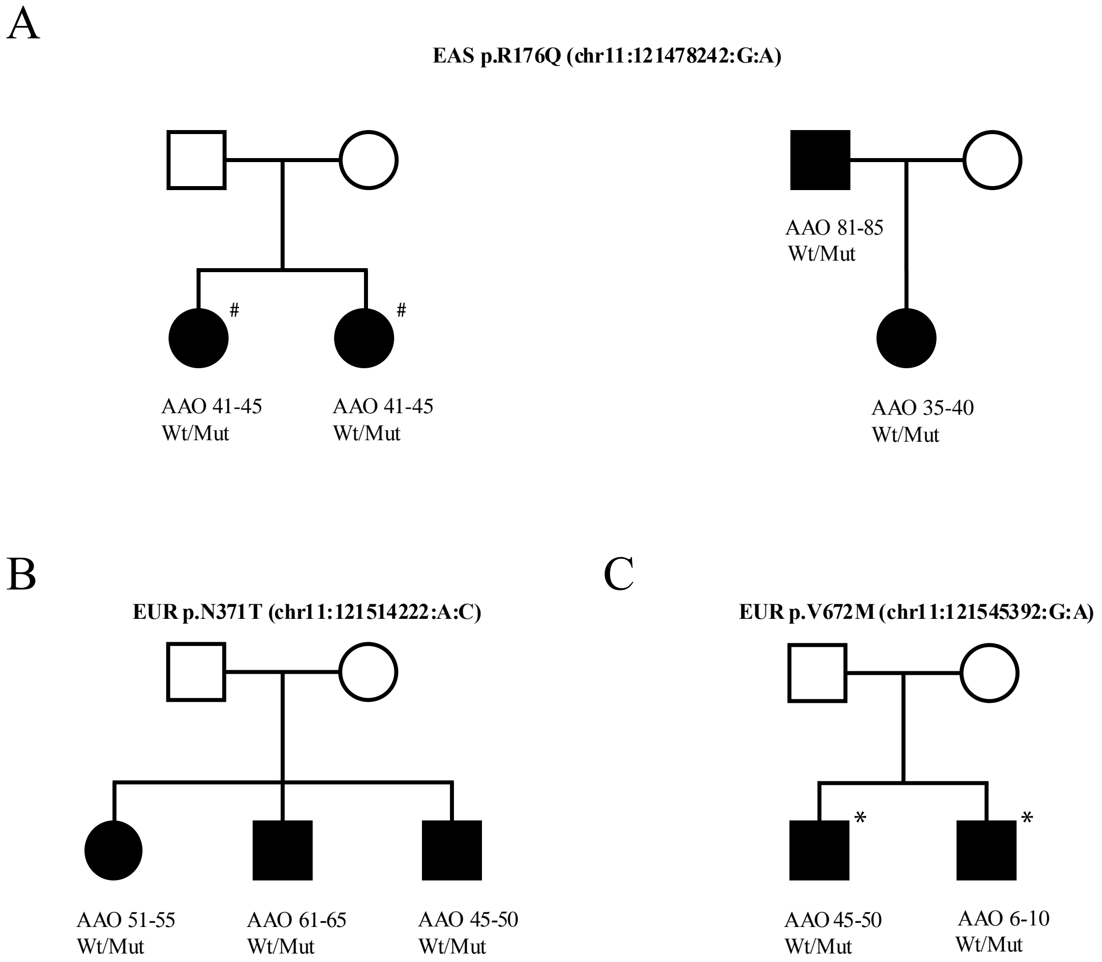
